# Detection of SARS-CoV-2 in Human Breast Milk

**DOI:** 10.1101/2020.04.28.20075523

**Authors:** Rüdiger Groß, Carina Conzelmann, Janis A. Müller, Steffen Stenger, Karin Steinhart, Frank Kirchhoff, Jan Münch

## Abstract

SARS-CoV-2 (CoV-2) is mainly transmitted in the human population during close contact and respiratory droplets. It is currently unclear, however, whether CoV-2 is shed into milk and may also be transmitted from infected mothers to newborns trough breast feeding. Two recent reviews on the topic (1,2) did not find evidence for CoV-2 in human milk. However, the number of breast milk samples analyzed so far is small and samples were taken only once from each mother (2).

---

It is currently unclear whether SARS-CoV-2 (CoV-2) may be shed into breast milk and transmitted through breastfeeding. Recent reviews and research articles reported no evidence for CoV-2 in human breast milk but the sample size was small.^1–3^

Here, we examined milk samples from two CoV-2 infected nursing mothers (M1 and M2). An overview on the clinical data and time course of infection is provided in Fig. 1A. We determined CoV-2 viral loads using RT-qPCR for CoV-2 N and -ORF1b^4^ (Fig. S1) in whole milk and skim milk obtained after removal of the lipid fraction. All four milk samples obtained from M1 from day 12 to 14 tested negative (Fig. 1B). In contrast, CoV-2 RNA was detected in milk provided by M2 on days 10 (left and right breast), 12, 13 and 14, while a later sample from day 25 was negative (Fig. 1A). Peak levels were detected on day 12 with Ct values for CoV-2 N of 29.8 and 30.4 in whole milk and skim milk, respectively (Fig. 1B, top), corresponding to viral genome copy numbers of 1.32 × 10^5^ and 9.48 × 10^4^ per ml (mean of both isolations, Fig. 1B, bottom). Since milk components may affect RNA isolation and quantification, viral RNA recovery rates upon spiking milk with serial dilutions of a CoV-2 stock were determined. We observed an up to 89.2% reduced recovery rate in whole milk and 51.5% in skim milk (Fig. S2) suggesting that the actual viral loads in whole milk of M2 may be even higher (Fig. 1C).

**Figure 1.**
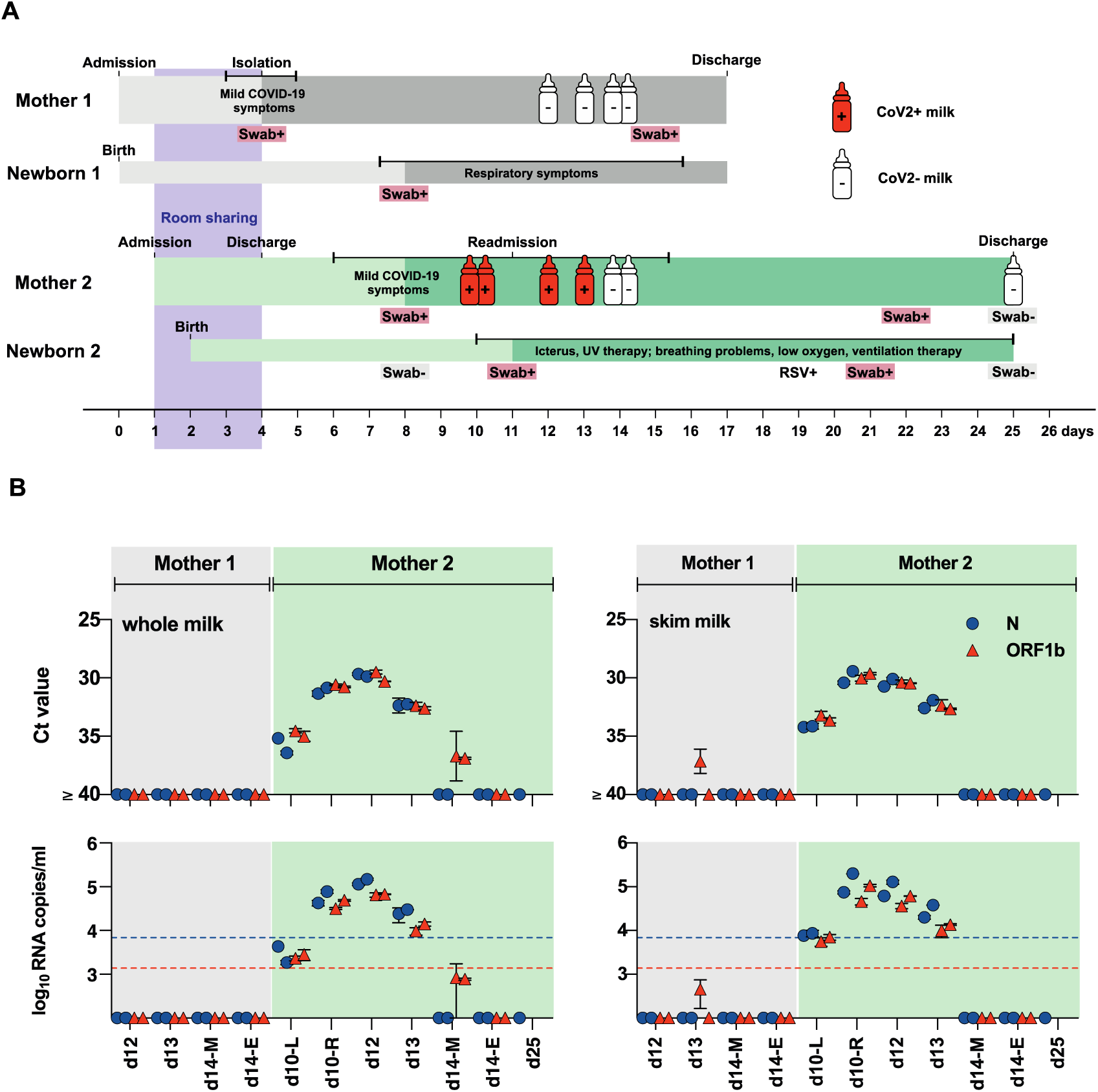
Detection of SARS-CoV-2 in milk of an infected mother. **A) Clinical data and time course of infection of both mothers with newborns**. After delivery, M1 developed mild COVID-19 symptoms and was subsequently tested positive at day 4 for CoV-2, resulting in spatial isolation of M1 with her newborn (N1). N1 was subsequently tested positive and developed respiratory problems, but both recovered and were discharged 18 days later. M2 was admitted to the same hospital and the same room as M1&N1. Upon delivery, M2 & N2 were brought back in the same room until M1 was diagnosed with CoV-2 and isolated. M2 & N2 were discharged at day 4 but shortly thereafter M2 developed mild COVID-19 symptoms (coughing, sniffing, low fever, loss of taste and smell) and wore since then a surgical mask for 24 hours per day. M2 was tested CoV-2 positive at day 8. At day 11, N2 was tested CoV-2 positive and readmitted to a specialized hospital because of newborn icterus. N2 then developed severe breathing problems including frequent decreases in oxygen levels requiring ventilation therapy and tube feeding, and then recovered. At day 19, N2 was tested positive for RSV and two days later again for CoV-2. M1 was tested CoV-2 positive again at day 22, 13 days after first being diagnosed with CoV-2. At day 26, M2 & N2 were tested negative. Dark shading indicates time from first CoV-2 positive swab **B) Detection of SARS-CoV-2 RNA in human milk samples by RT-qPCR**. RNA isolated from whole and skim milk of different timepoints for both mothers was subjected to analysis by RT-qPCR using primer sets targeting SARS-CoV-2-N and -ORF1b. Samples and viral RNA standard were run in duplicates, isolation and RT-qPCR repeated in two independent assays. RNA from milk of M2 at day 25 was only isolated once and only analyzed by CoV-N RT-qPCR. Symbols at baseline indicate no amplification (or Ct > 36.5 and no amplification in one replicate). Blue dashed line denotes quantification threshold for N (160 copies/rx, Ct 34.2) and red dotted line for ORF1b (32 copies/rx, Ct 35.9). Values below these lines but above baseline indicate amplification in both replicates, but no reliable quantification. Values shown are mean from duplicates ± SD. Top panel shows Ct values and bottom panel the corresponding viral RNA copy numbers/ml. Abbreviations: d, day; L, left; R, right; M, morning; E, evening

We detected CoV-2 RNA in milk samples collected over a period of 4 days. Based on specimen handling, internal controls, and negative testing of M1 milk, contamination can be excluded. Detection of viral RNA in milk of M2 coincided with mild COVID-19 symptoms and a CoV-2 positive diagnostic testing of the newborn (N2) (Fig. 1A). M2 was wearing a surgical mask since the onset of symptoms and followed safety precautions during handling or feeding the neonate, including proper hand and breast disinfection, strict washing and sterilization of milk pumps and tubes. However, whether N2 was infected by breastfeeding or other modes of transmission remains unclear. Further studies of milk samples from lactating women and possible virus transmission via breastfeeding are needed to develop recommendations on whether mothers with COVID-19 should breastfeed.

## Data Availability

All primary data will be made available to qualified individuals upon request.

## Authors’ Contributions

R.G., C.C. and J.A.M. contributed equally to this study. R.G. performed RNA extraction, RT-qPCR analysis and evaluated data. C.C. and J.A.M. generated virus stock handled milk samples and prepared them for RNA extraction. S.S. supervised BSL3 work and helped in writing. K.S. organized samples and was involved in study design. F.K. and J.M. supervised the project and wrote the article.

## Ethics statement

Both mothers were informed about the study and gave informed consent. Ethical approval for this study was waived by the Ethics Committee of Ulm University because this study is case report and all samples were anonymized.

## Notes

### Competing Interest Statement

The authors have declared no competing interest.

### Funding Statement

This research was supported by the European Union's Horizon 2020 project “Fight-nCoV” (to J.M.) in response to the corona virus outbreak for research and innovation action under grant agreement No.101003555. This research was also supported by the Collaborative Research Center 1279 by the German Research Foundation (to. S.S., F.K., and J.M.).

